# Bayesian Emulation and History Matching of JUNE

**DOI:** 10.1101/2022.02.21.22271249

**Authors:** Ian Vernon, Jonathan Owen, Joseph Aylett-Bullock, Carolina Cuesta-Lazaro, Jonathan Frawley, Arnau Quera-Bofarull, Aidan Sedgewick, Difu Shi, Henry Truong, Mark Turner, Joseph Walker, Tristan Caulfield, Kevin Fong, Frank Krauss

## Abstract

We analyse June : a detailed model of Covid-19 transmission with high spatial and demographic resolution, developed as part of the RAMP initiative. June requires substantial computational resources to evaluate, making model calibration and general uncertainty analysis extremely challenging. We describe and employ the Uncertainty Quantification approaches of Bayes linear emulation and history matching, to mimic the June model and to perform a global parameter search, hence identifying regions of parameter space that produce acceptable matches to observed data.

## 1 Introduction: Bayesian Emulation and Uncertainty Quantification

The Covid-19 pandemic disrupted healthcare systems and caused substantial fatalities around the globe. Various models have been developed to aid decision makers in the assessment of policy options, from simple analytic models up to complex agent-based models (ABMs). June, introduced in [3], is of the latter type, and its high level of demographic and spatial resolution demands substantial computational resources to evaluate. A critical component in the uncertainty analysis, and subsequent use for decision support of a complex epidemiological model such as June, is the process of model calibration: the matching of the model to observed data from the real system.

This process can be extremely challenging and in many cases its intractability precludes the full exploitation of sophisticated models, that may otherwise contain nuanced insights into the system of interest. The problem of calibrating a complex and computationally demanding model is not unique to epidemiology, and occurs in a wide range of scientific disciplines including cosmology, climate, systems biology, geology, energy systems [8, 46, 48, 49]. To solve this problem, an area of Bayesian statistics arose, sometimes referred to as the uncertainty analyses of complex computer models, or to use its more recent (and slightly more general name): the area of Uncertainty Quantification (UQ) [8, 19, 41]. UQ provides a statistical methodology combining a large number of efficient techniques with a set of overarching principles that address how to analyse complex models rigorously, transparently and robustly, for use in scientific investigations, for making real world predictions and for subsequent decision support. A full analysis of the behaviour of models with a large number of input parameters and possibly several outputs, and their subsequent calibration encounters the following three major issues:

1. For complex models, the evaluation time of the model is often so long that an exhaustive exploration of the model’s behaviour over its full input parameter space is infeasible.
2. When comparing models to observed real world data, an adequate statistical description of the link between model and reality, covering all major uncertainties and allowing for the rigorous use of an *imperfect* model, is required.
3. When calibrating, the appropriate scientific goal should be to identify *all* locations in input parameter space that lead to acceptable fits between model and observed data, and not just to find the location of a single good match.

We summarise in the next section three UQ methods: (a) Bayes linear emulation, (b) linking models to reality and (c) Bayesian history matching, that address the above three problems. We then apply these UQ methods to the June model in Sec. 3.

## 2 Bayesian Emulation and History Matching

### 2.1 Bayes Linear Emulation

Complex models typically have runtimes that can vary from seconds to days or even weeks, greatly inhibiting full model exploration, calibration, forecasting etc.. Many of the techniques in UQ therefore revolve around the construction of Bayesian *emulators*: statistical constructs that mimic the scientific model in question, providing predictions of the model outputs with associated uncertainty, at as yet unevaluated input parameter settings [36]. The emulators provide insight into the model’s core structure and, unlike the models they mimic, are extremely fast to evaluate, typically being several orders of magnitude faster. Hence they facilitate previously infeasible model exploration and global parameter searches. As an emulator makes predictions that have an associated (input dependent) uncertainty statement, they naturally fit within an overarching Bayesian uncertainty analysis, in which the impact of using an emulator instead of the model, can be understood and quantified. Emulators can be built for deterministic models, stochastic models, multilevel models (composed of models of increasing fidelity) and networks of models, providing a flexible and powerful set of tools to deal with a large class of scientific scenarios. Here we outline the construction of Bayes Linear emulators, a robust form of emulator, based on a partial specification, that has been successfully employed in several settings [8, 46].

We represent a general scientific model as the function *f* (*x*). Here *x* = (*x*_1_, …, *x*_*d*_) is a vector composed of all the input parameters. For example, *x*_1_ may represent an infectivity parameter, *x*_2_ a social distancing parameter etc.. *f* (*x*) = (*f*_1_(*x*), …, *f*_*q*_(*x*)) is the vector of all model outputs of interest, so for example *f*_1_(*x*) may represent the number of people hospitalised in England on a particular day, *f*_2_(*x*) the number of deaths on that day, all as a function of the inputs *x*. We anticipate that, due to limited computational resources, we will only be able to evaluate the model at a finite (and possibly small) number of input parameter locations *x*^(1)^, *x*^(2)^, …, *x*^(*n*)^ giving rise to model outputs *D*_*i*_ = (*f*_*i*_(*x*^(1)^), *f*_*i*_(*x*^(2)^), …, *f*_*i*_(*x*^(*n*)^))^*T*^, where *i* = 1, …, *q*. Therefore, at a new unevaluated input location, *x*, even say for a deterministic (i.e., repeatable) model, we will still be uncertain about the output value of *f* (*x*), as we will be for the vast majority of the input space. We take a subjective Bayesian view and treat the unknown *f* (*x*) as a random quantity, and construct an emulator that represents our beliefs about possible reasonable forms that this function *f* (*x*) could take. A popular emulator form for each output *f*_*i*_(*x*) is as follows [46]:

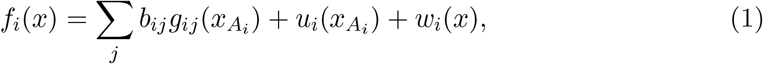

where we have selected a subset of the inputs, *x*, known as the active variables, 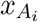, that are most influential for output *f*_*i*_(*x*). The first term on the right hand side of the Eq. (1) is a regression term, where *g*_*ij*_ are appropriately selected known deterministic functions of 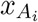, a common choice being low order polynomials, and *b*_*ij*_ are unknown scalar regression coefficients. The second term, 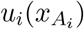, is a weakly second-order stationary process over 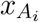, for which we only need specify its second order structure, choosing 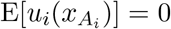, and utilising an appropriate covariance function: a classic example suitable for smooth functions is the squared exponential

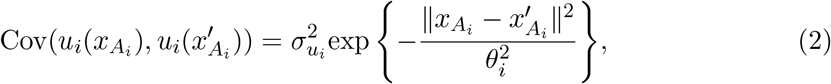

where 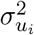 and *θ*_*i*_ are the variance and correlation length of 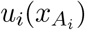 which must be specified a priori [46]. The third term, *w*_*i*_(*x*), is a white noise process uncorrelated with *b*_*ij*_, 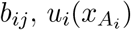, and itself such that

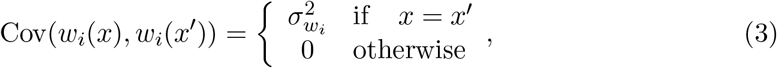

with expectation zero and 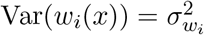. *w*_*i*_(*x*) represents the effects of the remaining inactive inputs not included in the list of 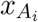, and formally facilitates a type of dimensional reduction [46].

The emulator form, as represented by Eq. (1), has various desirable features, and exploits our beliefs about the general anticipated behaviour of physically realistic models. The regression term, 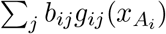, attempts to mimic the large scale global behaviour of the function *f*_*i*_(*x*): often substantial in physical models. The second term, 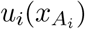, the weakly stationary process, mimics the local behaviour of *f*_*i*_(*x*), again exploiting concepts of smoothness of either *f*_*i*_(*x*) or attributes of *f*_*i*_(*x*) (if, say, *f*_*i*_(*x*) is stochastic). Such terms are highly versatile and can fit a large class of models, however, they require a sufficient density of runs to be suitably informed (regulated by the correlation length parameters *θ*_*i*_). In the literature there is sometimes an over-reliance on similar Gaussian Process (GP) style terms and a neglect of the regression terms, which may be unwise, as GPs of this form are typically capable of capturing the broad global behaviour, or the more complex local behaviour, but rarely both. We deliberately use the regression terms for the global structure, and utilise the 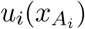 to capture the local behaviour.

We can select the list of active inputs, 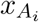, using various statistical techniques. For example, these could consist of classical linear model fitting criteria such as AIC or BIC [46], or automatic relevance determination [39]. A list of *p* active inputs for a particular physical output, *f*_*i*_(*x*), means that we have notably reduced the input dimensionality from *d* to *p*, which can result in large efficiency gains in subsequent calculations. The small remaining effect of the inactive inputs is not ignored, but is captured by the third term *w*_*i*_(*x*) in Eq. (1), whose variance 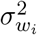 represents the added uncertainty induced by the dimensional reduction.

#### What to Emulate

A major issue when emulating complex models, is the choice of the set of attributes/outputs of the model to emulate. For example, often an objective function describing the mismatch between model and data has been emulated (e.g., a simple chi-squared metric, or a more complex likelihood function). However, despite being deceptively simple having just a single output, the objective function typically has a complex form, as it depends upon the union of all active inputs, and possesses numerous local maxima/minima [46], rendering this an inefficient strategy. Instead, we prefer to emulate the physical outputs of the model directly, as these tend to have a) a smaller list of active inputs per output allowing a nuanced and sometimes substantial dimensional reduction tailored to each individual output, and b) a simpler functional dependence on the input parameters that is often well represented by the regression terms in the emulator. Further choices are required when emulating stochastic models, where we can choose to emulate summaries of outputs of interest such as the mean, the variance, or quantiles if required, possibly conditioning on key events such as epidemic take-off, and extend for example to covariance structures between groups of outputs if needed. In these cases the role of *w*_*i*_(*x*) is extended to also incorporate the uncertainties induced by using estimates from finite samples [1, 2].

#### Designing batches of model evaluations

We begin by specifying the region of input space of interest, typically a *d*-dimensional hypercube, and denote this *χ*_0_ ⊂ ℝ^*d*^. We then design a set of ‘space filling’ runs over *χ*_0_, constructed to be well spread out, without any large holes. For example we may use a maximin Latin hypercube design, an approximately orthogonal design, also desirable for emulator construction [9, 40].

#### Updating the Emulator

We then update our prior emulator structure given by Eq. (1) with the information from the set of model runs using our favourite statistical tools. Specifically, we would prefer a fully probabilistic Bayesian approach if we required full probability distributions on all emulated outputs, *f*_*i*_(*x*) [18], and were we prepared to specify full joint probabilistic priors. However, in most cases our preferred choice is to use Bayes Linear methods, a more tractable version of Bayesian statistics which requires a far simpler prior specification and analysis [12, 14]. It deals purely with expectations, variances and covariances of all uncertain quantities of interest, and uses the following Bayes linear update equations, derived from foundational arguments [14], to adjust our beliefs in the light of new data. When emulating the *i*th output *f*_*i*_(*x*) of a complex model, where we had performed an initial batch of *n* runs giving a vector of model output values *D*_*i*_ = (*f*_*i*_(*x*^(1)^), *f*_*i*_(*x*^(2)^), …, *f*_*i*_(*x*^(*n*)^))^*T*^, we obtain the adjusted expectation, E_*D*_ (*f*_*i*_(*x*)), and adjusted variance, 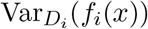, for *f*_*i*_(*x*) at new input point *x* using:

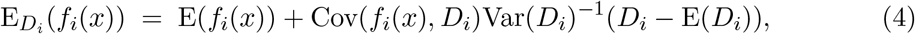

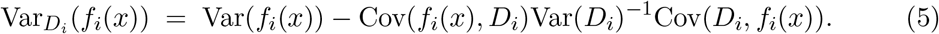

All quantities on the right hand side of Eqs. (4) and (5) can be calculated from Eqs. (1) and (2) combined with prior specifications for E(*b*_*ij*_), Var(*b*_*ij*_), 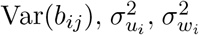 and *θ*_*i*_. 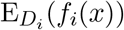 and 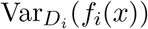 provide a prediction for *f*_*i*_(*x*) with associated uncertainty, and are used directly in the implausibility measures used for the global parameter searches described in Sec. 22.3. Note that multivariate versions of Eqs. 4 and 5 are available. In addition, we may make certain pragmatic choices in the emulator construction process, for example, while we typically keep the regression coefficients *b*_*ij*_ uncertain, we may directly specify 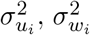 and *θ*_*i*_ a priori, or use suitable plugin estimates as described in [46]. We can test the emulators using a series of diagnostics, for example checking their prediction accuracy over a new batch of runs [5]. An example of a 1D emulator is given in Fig. 1, cf. [36] for an introduction and [18, 46, 47] for detailed descriptions of emulator construction.

**Figure 1:**
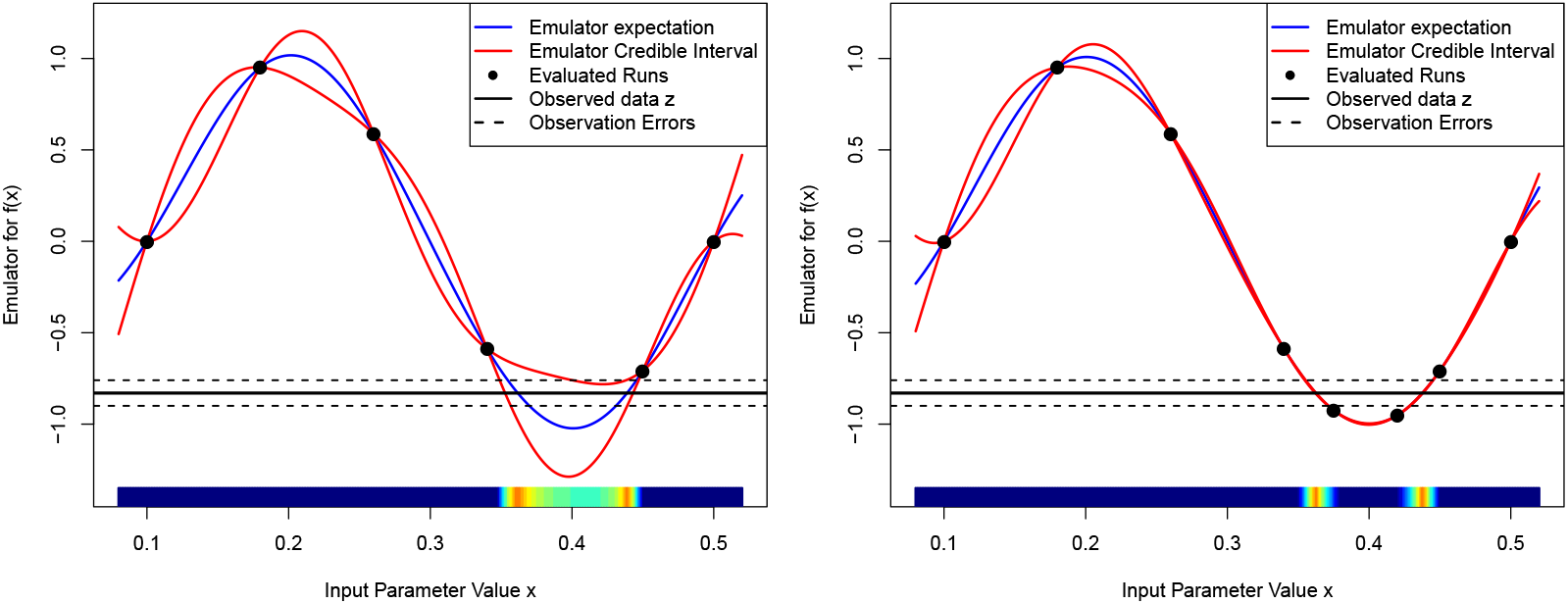
An emulator of a 1-dimensional toy model where *f* (*x*) = sin(2*π*(*x* − 0.1)*/*0.4), for the 1st wave/iteration, using just 6 runs (left panel), and for the 2nd wave, using 2 additional runs (right panel). The emulator’s expectation E_*D*_[*f* (*x*)] and credible intervals 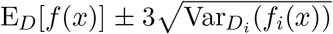 are given by the blue and red lines respectively, with the observed data *z* that we wish to match to as the black horizontal line (with errors). The implausibility *I*(*x*) is represented by the coloured bar along the *x*-axis, with dark blue implying *I*(*x*) *>* 3, light blue 2.5 *< I*(*x*) *<* 3 and yellow (*I*(*x*) *<* 1).

### 2.2 Assessing Uncertainties: Linking the Model to the Real World

Most epidemiology models are developed to help explain, understand and predict the behaviour of the corresponding real world system of interest, typically in terms of the progression through a population of an infectious disease. An essential part of determining whether such a model is adequate for this task is the comparison of the model with data formed from observations of the real system. However, this comparison involves several uncertainties that must be accounted for to provide a meaningful definition of an ‘acceptable’ match between a model run and the observed data. Hence it is vital to define a clear statistical model describing the difference between the epidemiological model, *f* (*x*), and the observed data denoted as the vector *z*. While more complex statistical models are available [13], here we describe a simple but powerful version that has been successfully used in a variety of scientific disciplines, for example climate, cosmology, oil reservoirs, epidemiology and systems biology [1, 8, 17, 46, 48, 49].

The most familiar source of uncertainty is observational or experimental. We represent the features of interest of the real system as a vector of uncertain quantities, *y*, which will be measured imperfectly involving a vector of errors *e*, to give the vector of observations, *z*, as

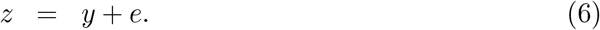

We represent the errors as additive here, but could use a more complex form if necessary. Depending on the scientific context, we then make judgements about the relationship between *y* and *e*, for example, a common specification [46] is to judge the errors *e* to be independent from *y*, with expectation, E(*e*) = *μ*_*e*_ and Var(*e*) = ∑_*e*_, a *q* × *q* covariance matrix. Setting *μ*_*e*_ = 0 corresponds to the judgement that the observations were unbiased, setting 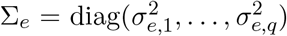, that is a diagonal matrix, corresponds to uncorrelated observation errors etc.

A critical feature that we must incorporate is the difference between the epidemiological model, *f* (*x*), of the system, and the real system, *y*, itself. We represent this difference between model and reality using a *structural model discrepancy* term. First we note that even were we to evaluate the model, *f* (*x*), at its best possible choice of input, *x*^*^, the output, *f* (*x*^*^), would still not be in agreement with the real epidemiological system value *y*, due to the many simplifications and approximations inherent to the model, therefore:

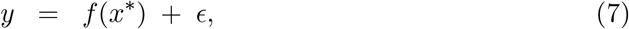

where *ϵ* is the structural model discrepancy: a vector of uncertain quantities that directly represents the difference between the model and the real system. Note that we are still treating *y, f, x*^*^ and *E* as vectors of random quantities. Now we have to make judgements about their relationships: a simple and popular specification [8, 46] would be to judge that *ϵ* is independent of *f* (*x*^*^), *x*^*^ and *e*, with E(*ϵ*) = 0. In the case of a single output we would then specify 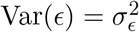. However, for the full case of *q* outputs, we may specify Var(*ϵ*) = ∑_*ϵ*_, a *q* × *q* covariance matrix. ∑_*ϵ*_ may have intricate structure possessing non-zero covariances between components of *E*, in order to capture the heavily correlated deficiencies of the model outputs. Various structures for ∑_*ϵ*_ of increasing complexity are available [8, 18, 46], along with methods for their specification [15, 46]. Note that typically the form of ∑_*ϵ*_ is very different from ∑_*e*_.

While the inclusion of the structural model discrepancy is unfamiliar to most modellers, it is now of standard practice in the Uncertainty Quantification literature for analysing complex but imperfect models [6, 7, 18, 47]. It facilitates a richer analysis whereby we can incorporate our necessarily uncertain knowledge of the model’s deficiencies to improve our modelling of reality *y*. Its inclusion can prevent over-fitting when calibrating and also reduces both bias and over-confidence when predicting. It is also vital when combining the results of several models.

### 2.3 Bayesian History Matching

Due to their fast evaluation speed, emulators can be used in a variety of UQ calculations that would be otherwise infeasible. One of the most important is the problem of performing global parameter searches. Here we outline a powerful iterative emulator based global search method known as History Matching (HM), which has been successfully employed in a variety of scientific disciplines [1, 8, 46, 49]. HM is designed to answer the questions:

1. Are there any input parameter settings that lead to acceptable matches between the model output and observed data?
2. If so, what is the full set X that contains all such input parameter settings?

Note the emphasis on finding *all* such acceptable matches: optimising to find a single good fit is not adequate for assessing the impact of parametric uncertainty, nor for making predictions.

HM proceeds iteratively by ruling out regions of input parameter space that can be discarded from further investigation based on *implausibility measures* [8]. For an unexplored input parameter, *x*, we can ask how far would the emulator’s expected value for the individual function output, *f*_*i*_(*x*), be from the corresponding observed value, *z*_*i*_, before we would deem it highly unlikely for *f*_*i*_(*x*) to give an acceptable match were we to actually evaluate the function at *x*. The implausibility measure, *I*_*i*_(*x*), captures this concept, and for an individual output is given by the distance 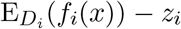 between emulator expectation and observed data, standardised by all relevant uncertainties,

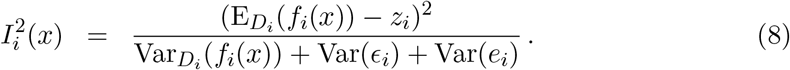

Here, 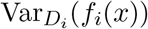 is the emulator variance, Var(*ϵ*_*i*_) the variance of the model discrepancy and Var(*e*_*i*_) the variance of the observational error, a direct consequence of Eqs. (6) and (7). See also Fig. 1 (the x-axis) for a depiction of *I*(*x*).

A large value of *I*_*i*_(*x*) for a particular *x* implies that we would be unlikely to obtain an acceptable match between *f*_*i*_(*x*) and *z*_*i*_ were we to run the model at *x*. Hence we can discard the input, *x*, from the parameter search if *I*_*i*_(*x*) *> c*, for some cutoff, *c*, which is often chosen by appealing to Pukelsheim’s 3-sigma rule [37], a very general and powerful result which states that for *any* continuous, unimodal distribution, 95% of its probability must lie within ±3*σ*, regardless of asymmetry or skew, suggesting that a choice of *c* = 3 may be reasonable [46]. This is the simplest univariate form, but we can combine implausibility measures from several outputs using say *I*_*M*_ (*x*) = max_*i*∈*Q*_ *I*_*i*_(*x*) for some set *Q*, or employ more complex multivariate forms [46].

Prior to performing the *k*th HM iteration, we define the current set of non-implausible input points as χ_*k*_ and the set of outputs that we considered for emulation in the previous wave as *Q*_*k*−1_. We proceed as follows [48]:

1. Design and evaluate a well chosen set of runs over the current non-implausible space χ_*k*_. e.g. using a maximin Latin hypercube with rejection [46].
2. Check if there are new, informative outputs that can now be emulated accurately (that were difficult to emulate in previous waves) and add them to the previous set *Q*_*k*−1_, to define *Q*_*k*_.
3. Use the runs to construct new, more accurate emulators defined only over the region χ_*k*_ for each output in *Q*_*k*_.
4. The implausibility measures *I*_*i*_(*x*), *i* ∈ *Q*_*k*_, are then recalculated over χ_*k*_, using the new emulators.
5. Cutoffs are imposed on the Implausibility measures *I*_*i*_(*x*) *< c* and this defines a new, smaller non-implausible volume χ_*k*+1_ which should satisfy χ ⊂ χ_*k*+1_ ⊂ χ_*k*_.
6. Unless a) the emulator variances for all outputs of interest are now small in comparison to the other sources of uncertainty due to the model discrepancy and observation errors, or b) the entire input space has been deemed implausible, return to step 1.
7. If 6 a) is true, generate as large a number as possible of acceptable runs from the final non-implausible volume χ, sampled depending on scientific goal.

The history matching approach is powerful for several reasons: (a) While reducing the volume of the non-implausible region, we expect the function *f* (*x*) to become smoother, and hence to be more accurately approximated by the regression part of the emulator, 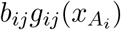. (b) At each new HM iteration we will have a higher density of points and hence the second term, 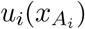, in the emulator should be more effective, as it depends on proximity to the nearest runs. (c) In later iterations the previously strongly dominant active inputs from early waves will have their effects curtailed, and hence it will be easier to select additional active inputs, unnoticed before. (d) There may be several outputs that may be difficult to emulate in early iterations (perhaps because of their erratic behaviour in uninteresting parts of the input space) but simple to emulate in later waves once we have restricted the input space to a much smaller and more epidemiologically realistic region. See [48] for further discussions comparing HM with Bayesian MCMC and ABC, and [21] for a direct comparison with ABC. We now apply these methods to the June model.

## 3 Application of Emulation and History Matching to JUNE

### 3.1 The June model

June [3] is an ABM that describes the spread of an infectious disease through large synthetic populations. Originally designed to simulate the circulation of Covid-19 through the English population, June has been also adapted to capture the populations of Cox’s Bazaar [4], a refugee camp in Bangladesh, and of Rhineland-Palatinate [42], one of Germany’s federal states. June’s description of the epidemic spread rests on four areas:

- the construction of a realistic synthetic population that reflects, as accurately as possible, the population demographic and their geographic distribution;
- the simulation of the population sociology, *i*.*e*. how the individuals behave: how they spend their time, whom they get into contact with and in which social environment;
- the parameterisation of the infection, how it is transmitted from infected to susceptible individuals and which impact it has on the health of infected individuals;
- the mitigation of spread and impact of the infection through pharmaceutical and non-pharmaceutical interventions (PIs and NPIs) such as social distancing and vaccinations, respectively.

They are discussed in more detail below.

#### 3.1.1 Population

June builds its synthetic population based on real or parameterised census data – in the case relevant for this contribution, June constructs the about 55 million residents of England based on the 2011 census data accessible through the NOMIS data base provided by the ONS. The data are organised hierarchically, with Output Areas (OAs) the smallest relevant unit, comprising on average about 250 residents with relatively similar socio-economic characteristics. The OAs have a specified geographic location and their data contain information about age, sex, and ethnicity of the area’s residents [26, 31, 33] and the composition of the households they live in [29], in about 20 categories^1^. June uses national averages to correlate age, sex, and ethnicity of individuals, which are presented as uncorrelated distributions in the data. In a similar way, information such as the national distributions of age differences of partners [30], and of parents and their children [24] are used to assign the individuals to their households.

As additional static properties of the population, June assigns school-age children to the nearest age-appropriate school; information about school locations and the age ranges for their students is taken from [10]. Within the schools, the students are grouped into class units of 20-30 individuals and have teachers assigned to them. In a similar way, Universities are filled with students – the young adults – and they are grouped into year groups of about 200 students.

The OAs are part of `Middle Super Output Areas` (MSOAs) with about 12500 residents, and 50 OAs constituting one MSOA. The census data provides information about the sectors of companies within MSOAs and about the distribution of the working population over these sectors, using the Standard Industrial Classification (SIC) scheme [34]. The parameterisation of company sizes with national sector-dependent averages allows June to construct an origin-destination matrix for the employees at the level of MSOAs [35]. Information concerning the commuting habits of individuals contained in the census data [32] underpins the construction of simplified virtual public transport networks within `JUNE`.

#### 3.1.2 Interactions

Having defined the static properties of the synthetic population – where people live, work and study – their daily lives outside work and education are filled with various activities. These activities include for example shopping, visiting friends and relatives in their homes, frequenting pubs and restaurants, going to the gym or cinema, to name a few, in the absence of any of these leisure activities, people are supposed to stay at home. Surveys performed *e*.*g*. by the Office for National Statistics [11] define the average proportion of time spent with various activities, in dependence on age and sex. These averages are translated into a probabilistic treatment thereby creating a highly flexible and varied daily schedule for `JUNE`’s virtual individuals.

These schedules are supplemented with contact matrices from `PolyMod` [22] and the BBC Pandemic Project [20], which indicate the average number of daily contacts – communication or physical – of individuals of age *i* with individuals of age *j* in different social settings ℒ, for example home (*H*), school (*S*), and work (*W*). As the contact numbers are presented as population averages, suitable for example for their deployment in compartment models, they need to be renormalized for the socially more granular IBMs^2^, resulting in the renormalized overall contact matrices 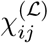 and the corresponding fraction of physical contacts, 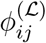, where ℒ ∈ {*S, H, W* }. While this introduces some uncertainty into the modelling of social interactions, the interplay of the synthetic population model with the contact matrices provides a welcome closure test for the self-consistency of the overall model.

For the purpose of fitting to data and the quantification of uncertainties in the model we assume that the construction of the synthetic population and its interactions is well understood and robustly and well parameterised as it is driven by data of relatively high quality,

#### 3.1.3 Infection

The description of the infection consists of two separate parts. First of all, the transmission from an infected person *i* to a susceptible person *s* needs to be simulated. In June as in many other models this is described as a probabilistic process. The infection probability for a susceptible person *s* with susceptibility *ψ*_*s*_ during a time interval from *t* to *t* + Δ*t*, spent with a group of individuals *g* in social context L is given as

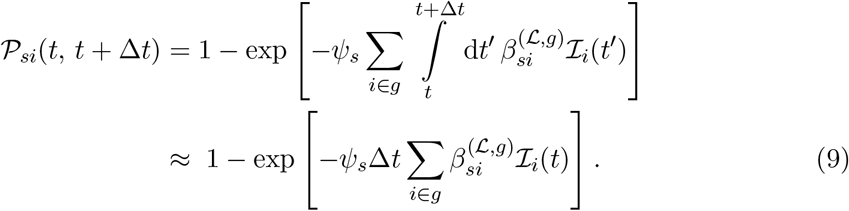

In the equation above, ℐ_*i*_(*t*) denotes the time-dependent infectiousness of the infected individual *i* in the group *g*. In June it follows a profile given by

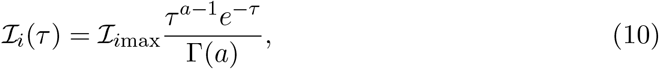

with *τ* = *t* − *t*_0_ − *t*_inc_, and *t*_0_ the time of infection of the individual, *t*_inc_ the incubation period and Γ the gamma function. *t*_inc_ is sampled form a normal distribution. The maximal or peak value of infectiousness for individual *i* is sampled from a log-normal distribution with median exp(*μ*) = 1 and shape parameter *σ* = 0.25, which allows for a long but small tail of highly infectious individuals which can be connected to superspreader events. The 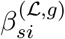 in Eq. (9) is the contact intensity between *s* and *i*,

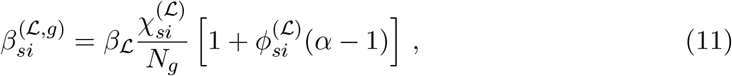

where the *β* are the social location-dependent baseline intensities, *N*_*g*_ is the number of individuals in the group setting, normalising the contact number *χ*_*si*_, and *α* parameterises the relative increase in infection probability for the proportion of physical contacts *φ*_*si*_. These parameters, the social-environment dependent *β* and the universal *α*, cannot be derived from first principles and must be obtained from fits to available data; they constitute a significant portion of the parameter space in the model and, correspondingly, a significant source of uncertainty.

Once an individual is infected, it takes some time – the incubation period – before they can infect others and some additional time before the onset of symptoms. A large range of input data has been used to derive various symptom trajectories for infected individuals, which in the case of high-income western countries in the global North depends mainly on their age and sex^3^. In the original formulation of the June model, significant efforts have gone into the quantification of probabilities for different health outcomes in the population, with some emphasis to also capture the health impact of Covid-19 on the highly vulnerable care home residents; we refer the reader to [3] for more details. Here, it should suffice to state that in June asymptomatic and symptomatic trajectories with varying severity have been identified, the latter ranging from mild, flulike symptoms over admission to regular or intensive-care wards to death in hospital or at residence. Although there are some uncertainties related to this treatment we usually do not consider them and treat the health outcomes as fixed by data.

#### 3.1.4 Interventions

Since the beginning of the Covid-19 epidemic, the UK government – like many other governments around the world – has employed a wide range of mitigation strategies. At the beginning of the pandemic, these interventions were mainly non-pharmaceutical, and these NPIs ranged from relatively simple strategies at the level of individuals, such as mask wearing and other social distance measures, to more involved and global strategies such as partial or complete lock-downs, involving school closures and the furloughing of parts of the work force. In June these measures can easily be modelled: social distancing measures and mask wearing can be described by modifying the *β*’s in the corresponding social settings by a factor, *M*_L_, capturing the reduced transmission probability, while the closure of schools or furloughing of the work force is easily described, based on data [16, 25, 45, 50], by keeping the impacted population at home instead of sending them to schools or work. For a more detailed description of the translation of NPIs to the June simulation we refer the reader to [3].

### 3.2 Inputs, Outputs and Initial Emulation

Our primary goal is to test if the June model can produce acceptable matches to observed data at the national and regional level, from the first wave of the Covid-19 pandemic and the subsequent summer period. We wish to identify the region of parameter space, χ, leading to such acceptable fits, if it exists. We identify a large set of input parameters, *x*, of interest to search over, primarily composed of interaction intensity parameters at the group level, and specify conservative ranges for each, given in table 1, which define the initial search space, χ_0_. A typical full England run of June would take approximately 10 hours to complete on 64 cores (Intel Xeon Skylake) and 128 GB of memory. This substantial computational expense combined with a relatively high dimensional input parameter space makes a global parameter search extremely challenging and necessitates the use of emulation. While there are several types of data available for the early pandemic, many of these had questions regarding reliability. For example, case data was substantially affected by the limited and evolving availability of Covid-19 tests, while hospital admission data was, especially during the peak of the first wave, collected with understandably varying levels of diligence across trusts. While it would in principle be possible to incorporate such data sets using a detailed statistical model for the measurement errors, *e*, in Eq. (6) that incorporated under-counting etc, we instead focus on hospital and total death data [27, 28, 43, 44], which although still uncertain due to the precise definition of death with Covid, suffers from far fewer issues.

**Table 1:**
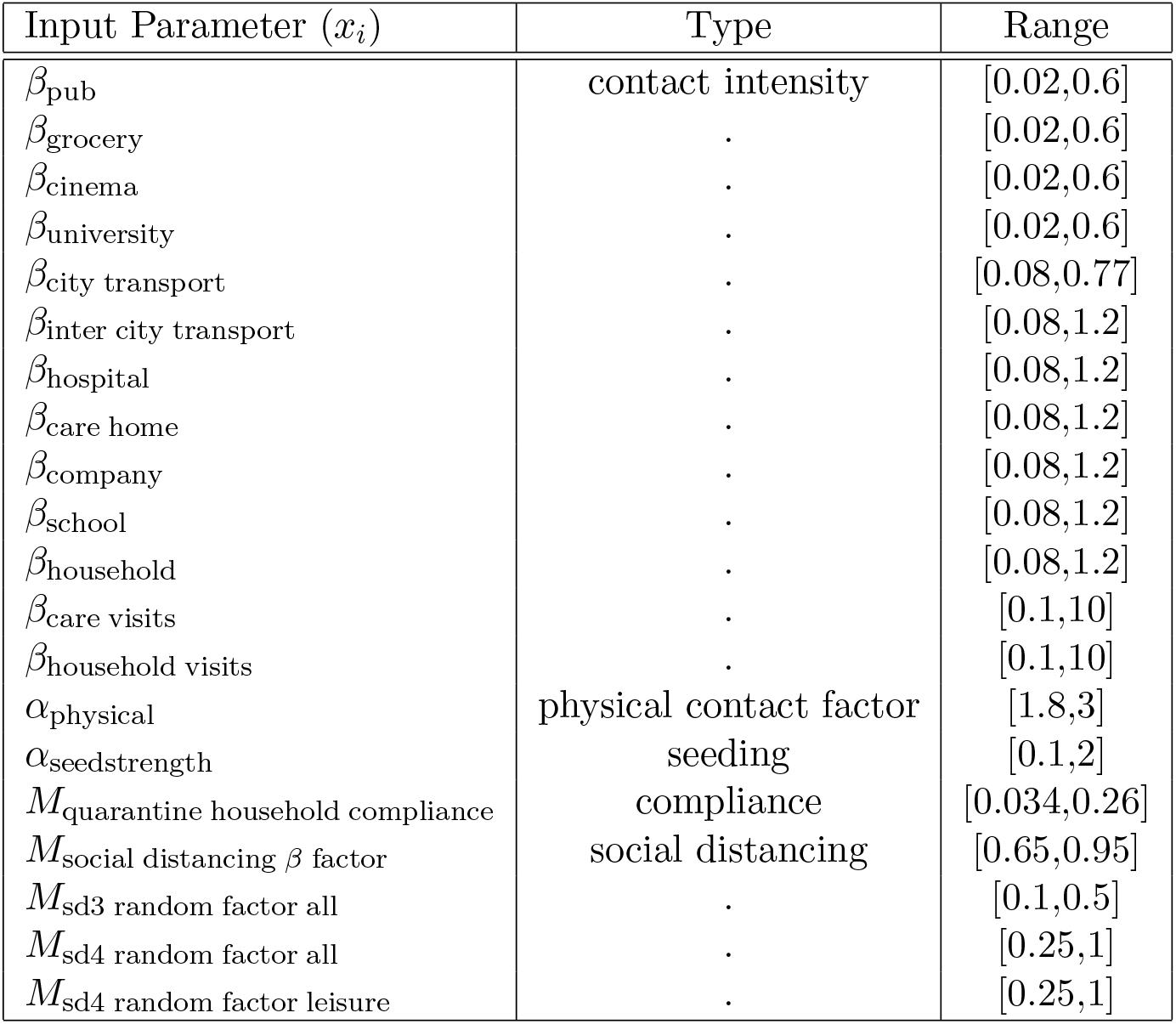
The input parameters explored in the global parameter search, their type and their ranges that define the search region *χ*_0_.

We define the primary June outputs of interest to be the hospital deaths and total deaths from 19th March to the end of August 2020, for England and its seven regions: East of England, London, Midlands, North East and Yorkshire, North West, South East, South West. We choose a subset of dates, shown as the vertical dashed lines in Fig. 4, to emulate. The observed data and June output is noisy, so we smooth them both using a standard kernel smoother (Gaussian kernel, bandwidth 7 days) as we wish to compare the underlying trends, and define the smoothed versions to be the target observed data points *z*_*i*_ (shown in Fig. 2), and the primary June outputs, *f*_*i*_(*x*). We specify conservative observation error and model discrepancy variances 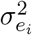 and 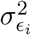 for each output as described in [3], by decomposing each into multiplicative and additive components to represent possible systematic biases, in addition to a scaled 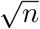 component for the observation error only, to model the noisy count process.

**Figure 2:**
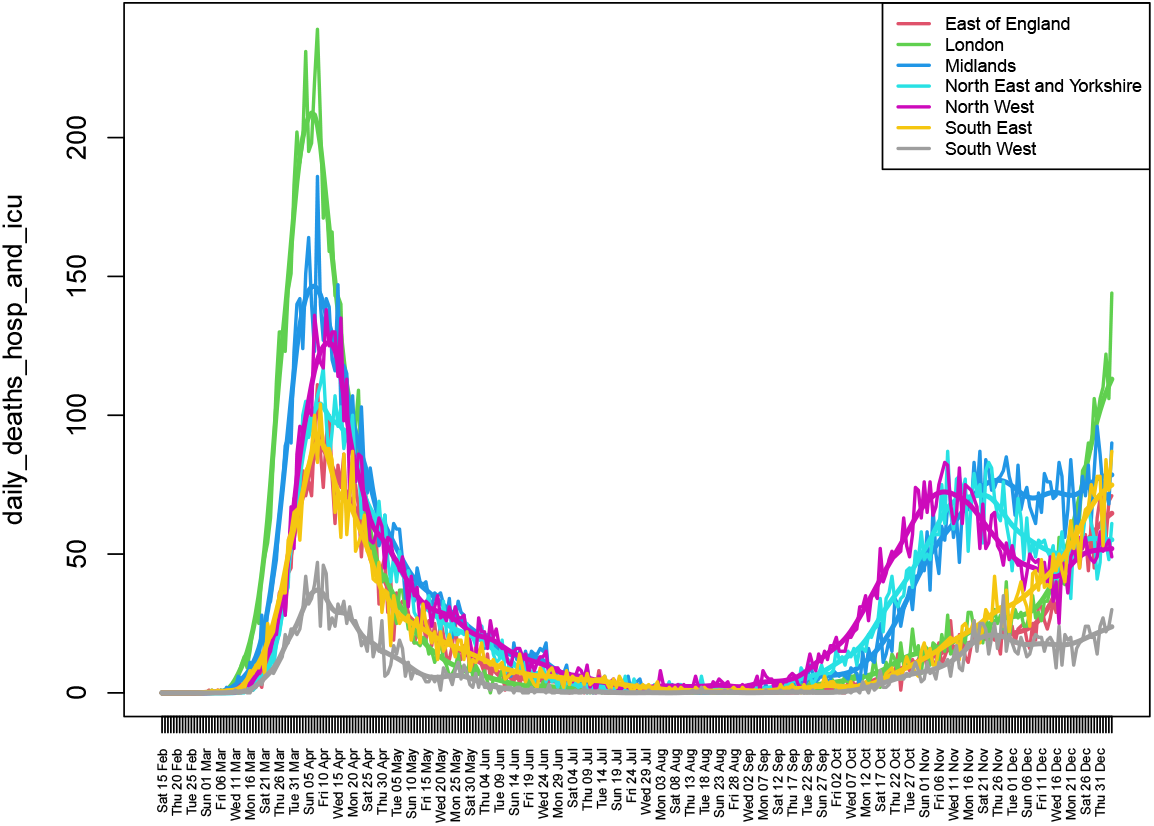
Daily Deaths in hospital wards and ICU in 2020, by region. The smoothed version used in the HM is also shown.

**Figure 3:**
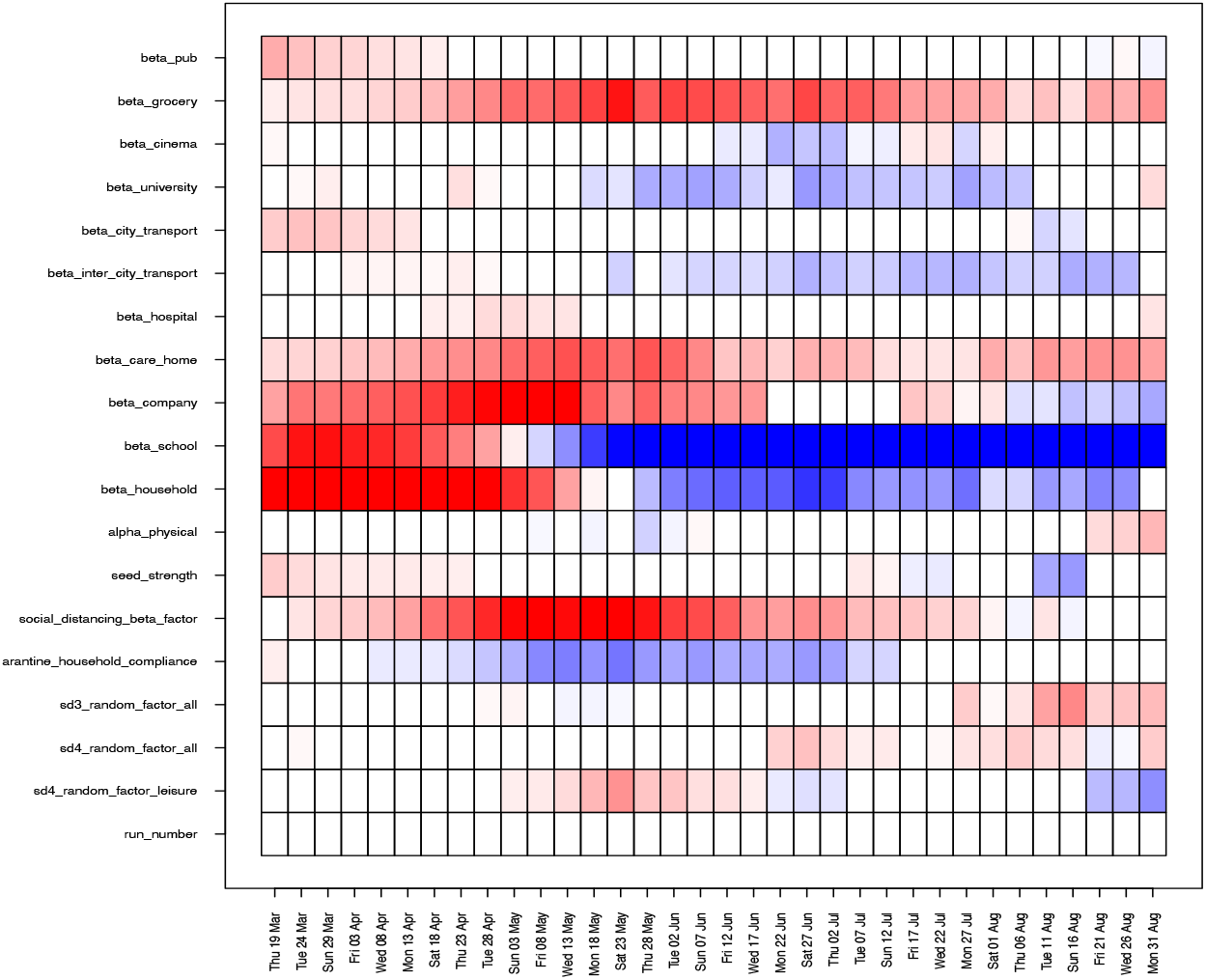
Estimates for the coefficients *b*_*ij*_ of the linear terms 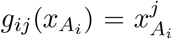 featuring in the emulator for total deaths in England, where *i* labels the time point (x-axis) and *j* labels the inputs (y-axis). Red/blue represents positive/negative dependencies of *f*_*i*_(*x*) on that input respectively, standardised as proportions of the largest coefficient for that time point.

**Figure 4:**
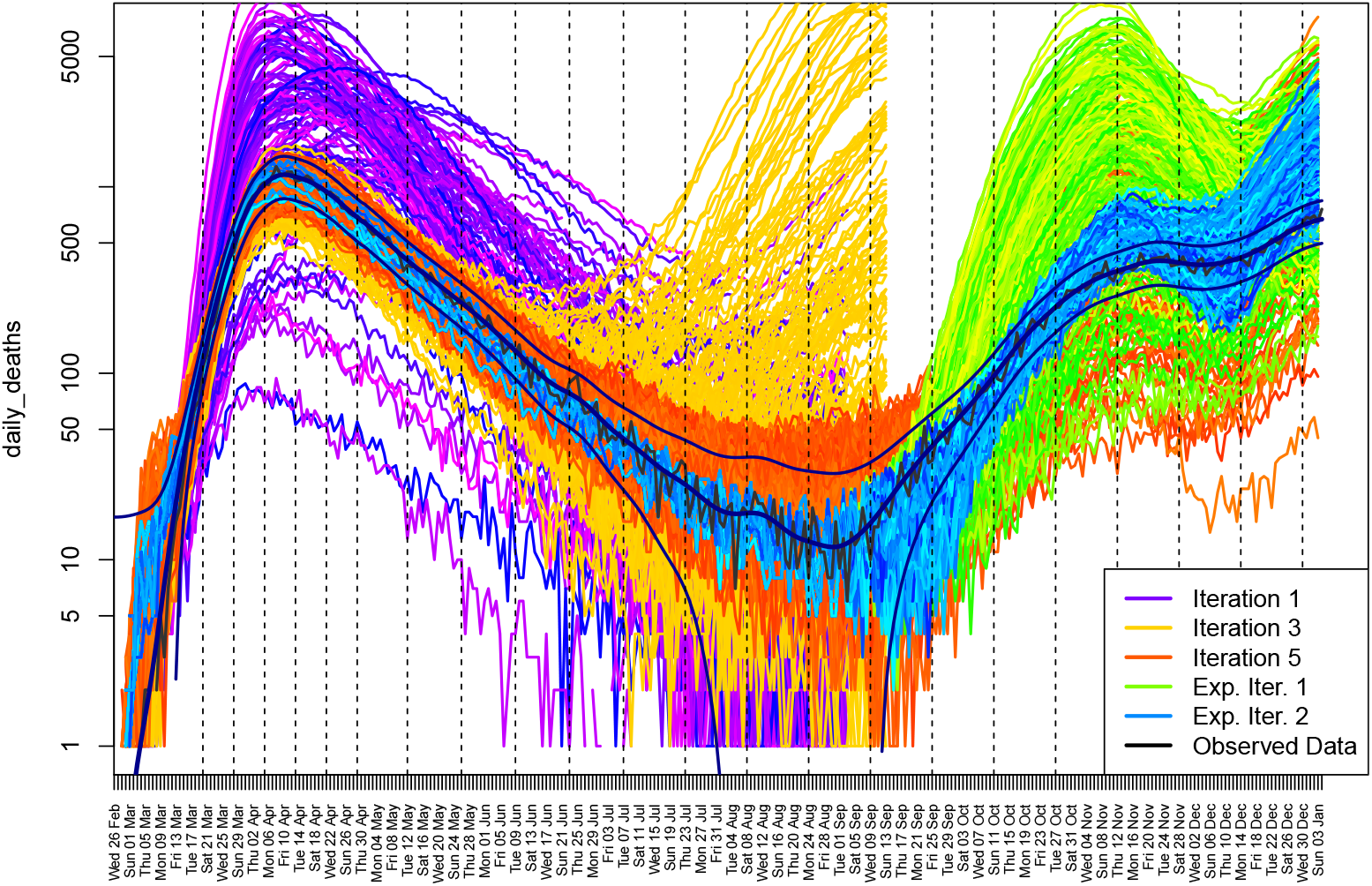
The June output for total daily deaths in England in 2020, for several iterations of the HM process. The smoothed and noisy data, along with the combined uncertainties due to *σ*_*e*_ and *σ*_*ϵ*_ are shown in black.

We design and evaluate a first wave of 150 runs over the 20-dimensional space, χ_0_, using a maximin Latin hypercube design. The outputs of these runs are shown as the purple lines in Fig. 4. We construct emulators for each output, *f*_*i*_(*x*), as detailed in Sec. 22.1 (using full quadratic regression terms selected using BIC, and MAP estimates for *θ*_*i*_ [46]). The emulators provide insight into the behaviour of the June model. For example, we can examine the coefficients *b*_*ij*_ of the linear terms 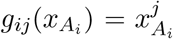 for the inputs featuring in Eq. 1, to gain insight into the effect each input has on each individual output. Estimates of these are shown in Fig. 3 for the total deaths outputs (each time point on the x-axis) with red/blue representing positive/negative dependencies respectively, standardised as proportions of the largest coefficient of that output. We see strong anticipated contributions from *β*_company_, *β*_school_ and *β*_household_ in the first wave of the pandemic from March to May, and more modest effects from *M*_social distancing *β* factor_ and *β*_grocery_ throughout the summer period. The sensitivities of *β*_school_ and *β*_household_ change to negative (blue) by May, as at this stage in many runs herd immunity has been reached, and hence increasing *β*_school_ will decrease deaths (as they will be brought forward in time). More insight can be gained from full emulator sensitivity analysis [23].

### 3.3 Iterative History Matching

We now employ the History Matching framework from Sec. 22.3, iteratively removing parameter space based on current implausibility measures, and performing batches/iterations of further runs. Fig. 4 shows the outputs from iterations 1, 3 and 5 for total deaths in England as the purple, yellow and red lines. As the iterations proceed the emulators become more accurate, we learn more about the global parameter space, and hence the runs approach the observed data, yielding reasonable matches across the first Covid-19 wave. The region of 20-dimensional parameter space deemed non-implausible at iteration 5 is shown in Fig. 5 as an *optical depth plot* which simply shows the depth in the remaining 18 dimensions of the non-implausible region (see [48]). Fig. 5 gives insights into the constraints imposed on the parameters by the death data and corresponding uncertainty specification. For example we see that we learn a lot about certain influential parameters such as *β*_school_ which are fairly well constrained while others such as *β*_care home_ can take a wider range of values, which we subsequently further constrained by specifically adding deaths in care home settings to the calibration effort^4^. We also see interesting relationships between pairs of parameters e.g. the reciprocal relations between *β*_company_ vs. *β*_household_ suggesting one or other can be high, but not both. We see similar relations between *β*_company_ vs. *M*_social distancing *β* factor_. Note that in using HM in this way, we do not seek to probabilize the non-implausible region as in a full Bayesian calibration, but we could go on to do this (*e*.*g*. by routing the emulators through an MCMC algorithm) if desired, but the additional information gained may be in part an artefact of the particular additional distributional choices that such an analysis requires, which may impact robustness.

**Figure 5:**
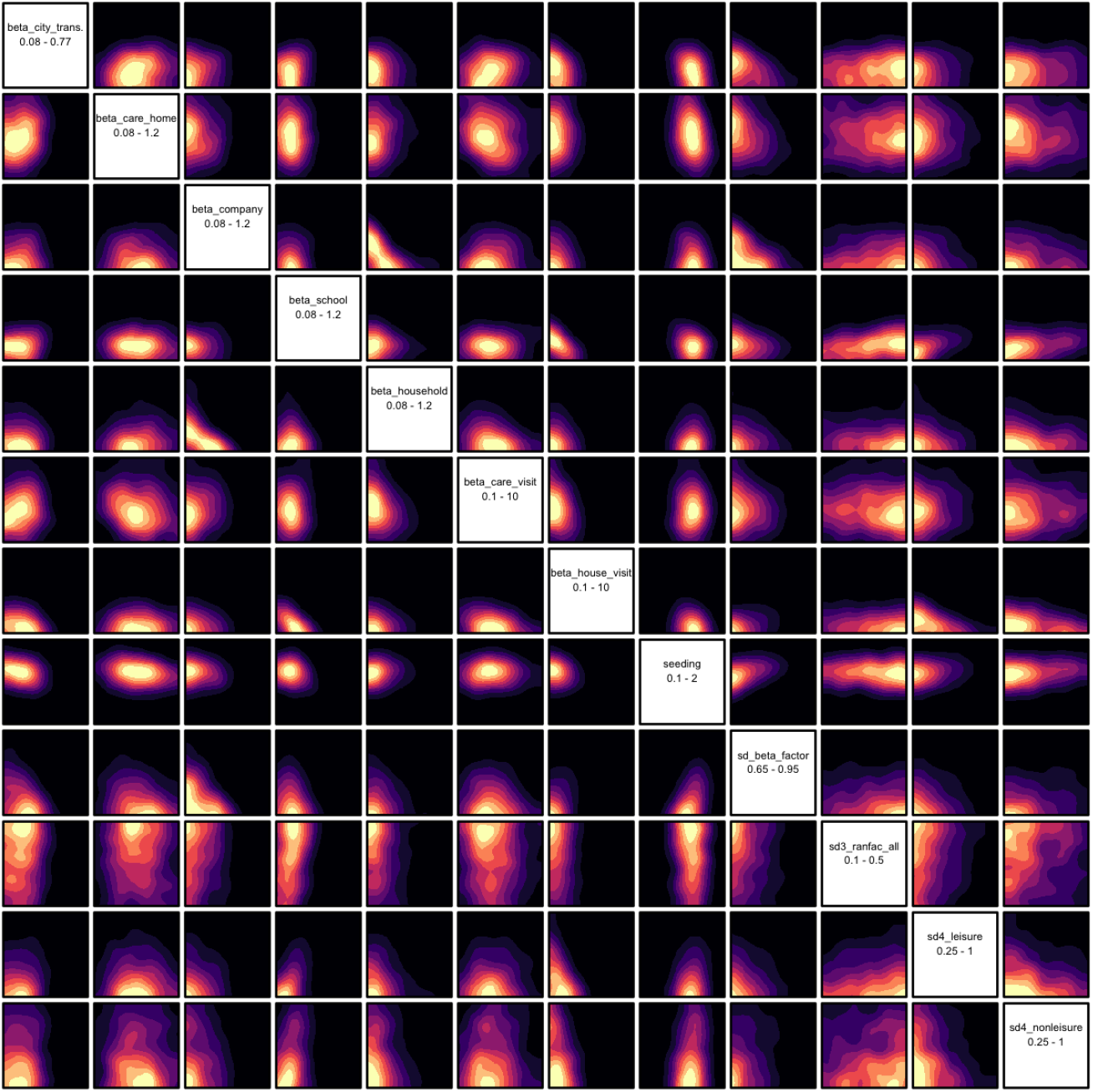
The optical depth of various 2-dimensional projections of the full 20- dimensional non-implausible region X_5_ found after the 5th iteration, with yellow implying greater depth. This region corresponds to the red runs in Fig. 4.

While performing a full global exploration of the input parameter space is of course preferable, it is sometimes useful to perform a fast “look-ahead” stage to check if such an expensive model is capable of fitting the next period of observed data, or whether model improvements are required. Fig. 4 also shows the results of such an exercise, where we took 8 runs with acceptable matches to the death data up to the end of August, and performed small 30-point designs for each of the 8 cases up until Dec 2020, now varying only 5 additional parameters relevant to the second Covid-19 wave (social distancing for schools, leisure, and non-leisure activities, Nov lockdown and B117 variant infectiousness), the output of which is given by the green lines. One iteration of HM was performed to reduce the 5-dimensional parameter space in each case, and a new set of runs designed which are shown as the blue lines in Fig. 4. We see reasonable matches to the first part of the second Covid-19 wave, with perhaps a late take-off in early September, and a partial overshoot in Nov-Dec, suggesting that June may well provide acceptable matches after a full HM. The earlier take-off of the data at the start of September is a particularly interesting effect as it is too early to be caused by schools opening. Even though we do include in our model the “eat-out-to-help-out” scheme which could help explain the infection uptick, we do not consider the effect of people movement due to holiday periods, which may be relevant here.

To give more detail, Fig. 6 shows a single unsmoothed run (red lines), from this final batch, but now for hospital deaths and total deaths for England and all seven regions, and shows the sort of quality of matches we are seeing so far. The black points give the (unsmoothed) death data and the combined uncertainties due to *σ*_*e*_ and *σ*_*E*_ shown as the blue lines.

**Figure 6:**
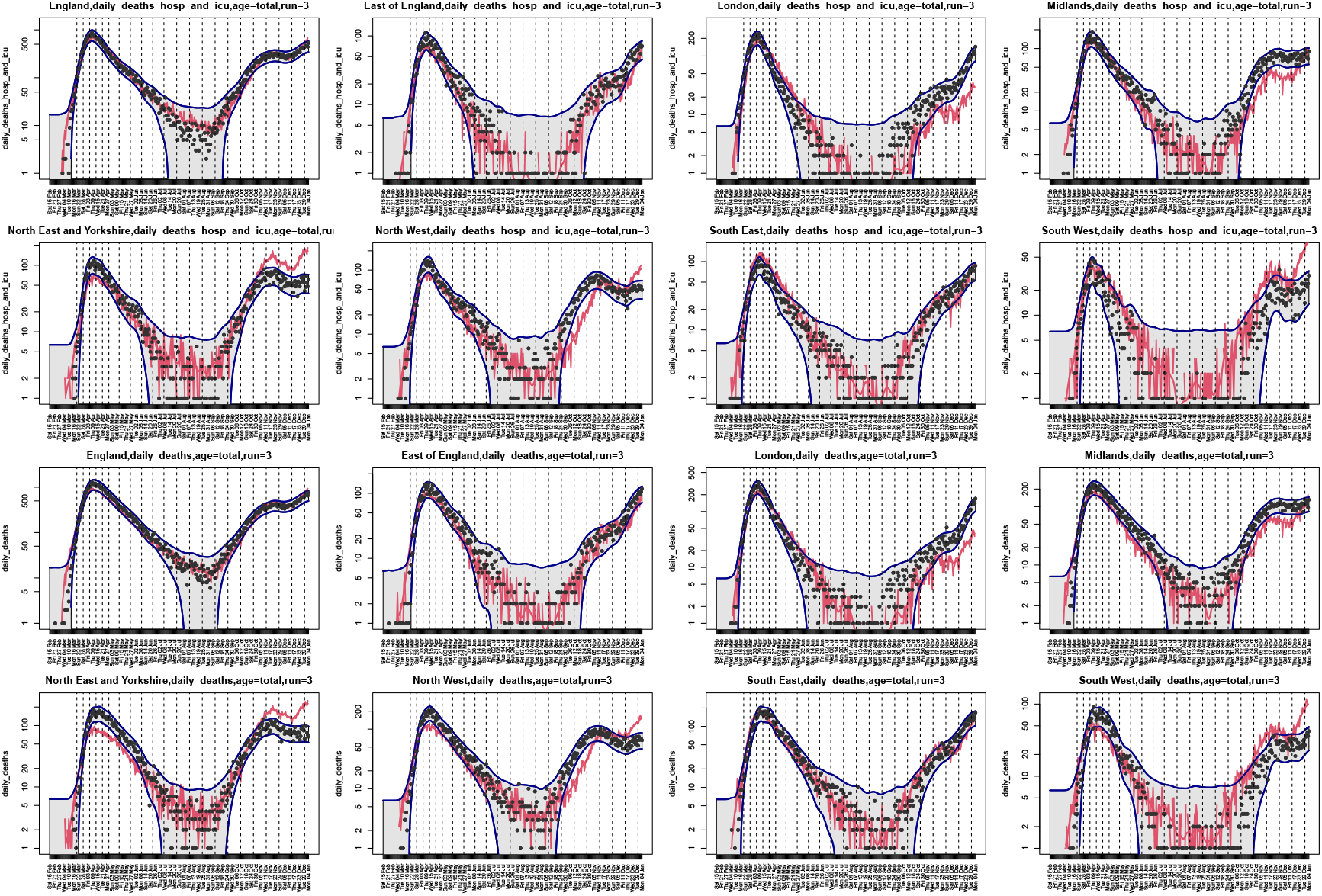
A single June run (red lines), from the second exploratory iteration (i.e. one of the blue lines in Fig. 4). The panels show hospital deaths (rows 1 and 2) and total deaths (rows 3 and 4) for England and the seven regions, as given in the plot titles. The black points give the (unsmoothed) death data, and the combined uncertainties due to *σ*_*e*_ and *σ*_*ϵ*_ are shown as the blue lines.

### 3.4 Discussion

Models such as June, with its high level of demographic and spatial granularity may become important tools to aid local and national decision makers. However, to fully exploit the nuances of such complex and expensive ABMs, efficient and comprehensive calibration methods are required. We demonstrated the emulation of June, providing insight into the model structure, and employed HM to identify the region of parameter space yielding reasonable matches to national and regional level hospital and total death data for the first Covid-19 wave. Such techniques form an essential tool for the future use of complex epidemiological ABMs, expanding our capabilities to combine detailed models with rigorous UQ. The ability to perform global exploration of the parameter spaces of expensive models of this form and to embed this within a broader UQ framework, is vital for making predictions with realistic uncertainty statements, and hence vital for subsequent decision support.

## 4 Outlook/Future Directions

Our work represents an important step towards the full exploitation of highly granular and detailed ABMs in health settings and elsewhere, harnessing the full depth of their simulations in providing high-quality understanding of critical dynamics and robust quantitative projections for improved decision support.

The next steps in this project are to include further outputs of interest within the HM for June, (hospitalisations, case rates, age categories etc.) and to examine smaller geographic regions, in which the stochasticity of June will become more pronounced, as compared to the national/regional level where it is somewhat subdominant. This will require more sophisticated emulator strategies [2], and if we are interested in detailed spatial predictions, will require the updating of the June state vector using UQ style data-augmentation techniques. Beyond this, these UQ methods are currently being incorporated wherever June is being employed e.g. by the UN for Cox’s Bazaar [4], a refugee camp in Bangladesh, and for Rhineland-Palatinate [42], one of Germany’s federal states.

In addition we plan to use the model to investigate in more detail social imbalances in COVID attack rates and infection-fatality ratios, which are relatively easy to trace in a model such as June. Supplementing the model with the elaborate UQ techniques will allow us to identify, in more detail and with increased certainty, important correlations between socio-economic markers of the population and the infection dynamics and outcomes.

## Data Availability

All data produced in the present study are available upon reasonable request to the authors.

## Ethics

No ethical issues.

## Data Accessibility

A full open source code base and implementation examples are available through

- github: https://github.com/IDAS-Durham/JUNE, and
- pypi: https://pypi.org/project/june/.

The version of June used for this work was v1.0 [38].

## Author’s Contribution

I.V. and J.O. created the Bayesian Emulation, History Matching framework for Uncertainty Quantification and calibrated the model, J.A.B., C.C.L., and A.Q.B. designed and implemented the June model, J.F. and M.T. maintain, document, and further improved the code base and its performance, A.S., D.S., H.T., and J.W. ran the code on various platforms and compared its results with relevant data, T.C. and K.F. provided valuable front-line insights from the health sector and data, F.K. initiated the creation of the June model and led its design, I.V. and F.K. wrote the paper. All authors read and approved the manuscript.

## Acknowledgements

This work was undertaken as a contribution to the Rapid Assistance in Modelling the Pandemic (RAMP) initiative, coordinated by the Royal Society. We are indebted to a number of people who shared their insights into various aspects of the project with us: We would like to thank Sinclair Sutherland for his patience and support in using the ONS database of the census data. We are grateful to Bryan Lawrence, Grenville Lister, Sadie Bartholomew and Valeriu Predoi from the National Centre of Atmospheric Science and the University of Reading for assistance in improving the computational performance of the model. The authors are grateful for inspiring collaboration with Richard Bower, Aoife Curran, Edward Elliot, Miguel Icaza-Lizaola, and Julian Williams in initial phases of the project.

## Footnotes

1 The English and Welsh census distinguishes between children, young (dependent) adults, such as University students, independent adults and old adults and classifies households according to the respective numbers in them.

2 As an example consider the number of contact children have with adults in schools. Clearly the number of contacts of average adults with children in schools is much less than the number of contacts adult teachers have with the children, necessitating a renormalization of the number of contacts by the proportion of teachers in the overall adult population.

3 In other population settings, for example in refugee camps, age and sex do not constitute good proxies for the overall health of an individual, and co-morbidities need to be explicitly factored in, giving rise to significant additional assumptions and uncertainties.

4 To the best of our knowledge this has not been yet tried or achieved by any other model.

